# Estimating Within- and Between-Family Polygenic Effects For Psychiatric Disorders Under Non-random Ascertainment

**DOI:** 10.64898/2026.07.28.26359090

**Authors:** Tahereh Gholipourshahraki, Ciarrah-Jane Shannon Barry, Oleguer Plana-Ripoll, Cynthia M. Bulik, Michael Eriksen Benros, Preben Bo Mortensen, Esben Agerbo, Liselotte Vogdrup Petersen, Bjarni Jóhann Vilhjálmsson

## Abstract

**Background:** Polygenic scores (PGSs) are increasingly used to investigate the genetic architecture of complex traits. In genetics, family study designs are often used to adjust for confounders such as population structure and shared environment. However, family studies may also be particularly vulnerable to to non-random ascertainment, for example when individual case status affects the probability of inclusion, leading to differential representation of sibling pairs. In sibling samples, PGS associations can be decomposed into within-family and between-family components, where the within-family estimate captures associations between sibling differences in PGS and differences in outcome, thereby providing an estimate that is less affected by shared familial confounding.

In this study, we examined the impact of non-random sampling on estimated genetic effects in family-based studies using both simulations and real-world data. Further, we leveraged the iPSYCH study design to estimate within-family PGS effects for six common mental health outcomes and whether accounting for these can improve prediction accuracy.

**Methods:** We conducted simulations and applied the same framework to real-world data to evaluate the impact of selection bias on within- and between-family PGS estimates. Selection bias was modelled through differential sampling of sibling pairs based on case status, and inverse probability weighting (IPW) was applied to adjust for known heterogeneous inclusion probabilities. Analyses were replicated in the iPSYCH cohort using registry-based sampling weights and PGSs for six major psychiatric disorders. Predictive performance of models was assessed using five-fold cross-validation.

**Results:** In simulation studies, biased sampling led to deviations in estimated PGS effects, with greater distortion observed for between-family components. IPW adjustment reduced the discrepancy between estimates obtained from biased and true underlying data. In the iPSYCH cohort, between-family estimates from unweighted models were larger than within-family estimates across traits. IPW weighted attenuated several of these estimates. Prediction analyses comparing models using total PGS versus decomposed within- and between-family components showed minimal differences in area under the curve and scaled R^2^ in the iPSYCH data, while modest gains were observed in selected simulation scenarios.

**Conclusions:** Non-random ascertainment distorts effect estimates in family-based models, with particular sensitivity when estimating between-family effects. Incorporating IPWs derived from known or estimable inclusion probabilities can reduce this bias. Our findings highlight the importance of accounting for selection bias in family studies when estimating genetic effects

## Introduction

The advent of large population-scale genetic data has over the last decade transformed research of complex traits, enabling genome-wide association studies (GWASs) to identify statistical associations between common genetic variants and phenotypic variation ^1-3^. Although single variants typically explain only a very small proportion of trait variance, their effect size estimates can be combined to construct polygenic scores (PGS). These summarise an individual’s genetic predisposition for a given phenotype ^4^. Increasing GWAS sample sizes have improved the predictive performance of PGS, encouraging widespread use in behavioral, biomedical and social research domains, with potential application in risk stratification and targeted prevention ^5-7^.

Family-based study designs, such as extended pedigrees or sibling-based analysis, have long been used in human genetics prior to the rise of population-based GWAS. Their renewed use reflects growing interest in distinguishing direct genetic effects from confounding factors such as assortative mating, genetic nurture, and population structure. Family-based study designs differ from other designs in their ability to account for shared environmental and genetic factors. Designs exploiting within-family segregation primarily provide estimates robust to gene–environment correlation (rGE) and population stratification. Within-family estimates may therefore provide a more accurate quantification of direct genetic effects to construct polygenic predictors that better reflect inherited liability, rather than correlated environmental pathways. Recent methodological developments further demonstrate incorporating relatives can increase statistical power while maintaining protection against confounding. Further, analyses not possible in samples of unrelated individuals, such as assessing parental origin effects and distinguishing inherited from non-inherited alleles, may be performed in within-family designs ^8, 9^.

The random segregation of parental alleles in sibling comparison models ensures genetic differences between siblings are independent of shared environmental influences, allowing effect estimates to be less affected by rGE and population structure ^9-13^. Several recent studies have demonstrated effect size estimates from unrelated individuals are often smaller when re-estimated within sibling pairs, evidencing the need to address familial confounding to obtain accurate PGS associations ^10, 14, 15^. These developments have contributed to the growing perception that family-based analyses offer a more accurate study of genetic influences, environmentally correlated exposures and evaluate PGS-trait associations.

However, family studies may still be affected by selection bias, particularly among families with genotyped siblings. Selection bias can arise from multiple sources, such as volunteer bias, attrition bias, non-response bias and sampling bias. As genetic studies typically only include individuals who have agreed to provide biospecimens for research, patterns of participation that may lead to non-representative inclusion of families are induced. Such selection bias is highly relevant when considering the potential clinical utility of PGS, as it may compromise population representativeness ^16, 17^. Inverse probability weighting (IPW) is a well-established method for addressing such bias ^18^. A weight is assigned to each individual, derived from their inverse probability of sample inclusion. Limited information about sample non-participants may be available in literature or census data. However, biobanks integrated with healthcare systems uniquely enable utilisation of existing demographic and clinical data to investigate factors influencing selection probability.

Several studies have employed family-based designs using sibling data to examine PGS-complex trait relationships ^10, 14, 15, 19^. Most current applications use the total PGS derived from unrelated individuals. If part of the between-family association is driven by confounding or selection processes, prediction based on total PGS may be inflated. Decomposing the score could provide more reliable prediction by focusing on direct genetic effects. Evidence on whether this approach enhances prediction in real-world data remains limited, particularly in the presence of non-random sampling. To our knowledge, no prior studies have investigated or adjusted for selection bias in this context. In the present study, we simulated sibling genetic data and considered multiple differing scenarios of non-random sampling. We evaluated the performance of IPW to correct for selection bias in each simulated scenario. Subsequently, we applied this correction to real-world data, using genotyped sibling pairs from the iPSYCH cohort ^20^. We leveraged nationwide register data from the full base population to estimate the probability of inclusion for each sibling pair, then used these probabilities to calculate IPWs. These weights were incorporated into within-family analyses and PGS associations were re-estimated to assess the impact of selection on PGS associations. Furthermore, we compared the predictive performance of models using total PGS with models including decomposed within- and between-family components to evaluate whether this framework improves prediction accuracy in simulated and real-world data.

## Results

Figure 1 provides a schematic overview of the analytic workflow implemented in this study. To investigate the impact of selection bias in family-based genetic association studies, we applied within- and between-family PGS models to both simulated and real-world data. These models decompose the total PGS association into two components; the within-family effect, to reflect the association between deviations from the family mean PGS and phenotype, and the between-family effect, to capture the association between the family average PGS and outcome. In our simulation study, we generated genotypes for sibling pairs and simulated binary phenotypes under a liability threshold model with known heritability and causal architecture. We oversampled case-control families to simulate selection bias and tested the use of IPW as a correction of this bias. In real-world data analyses, we used full sibling genotype and registry data from the iPSYCH cohort ^20^. PGSs were derived from large-scale GWAS summary statistics for major psychiatric traits, and sampling weights were constructed based on known inclusion probabilities ^21^. Logistic regression models were fitted to assess within- and between-family associations both with and without IPW adjustment. Further methodological details are provided in **Methods**.

**Figure 1.**
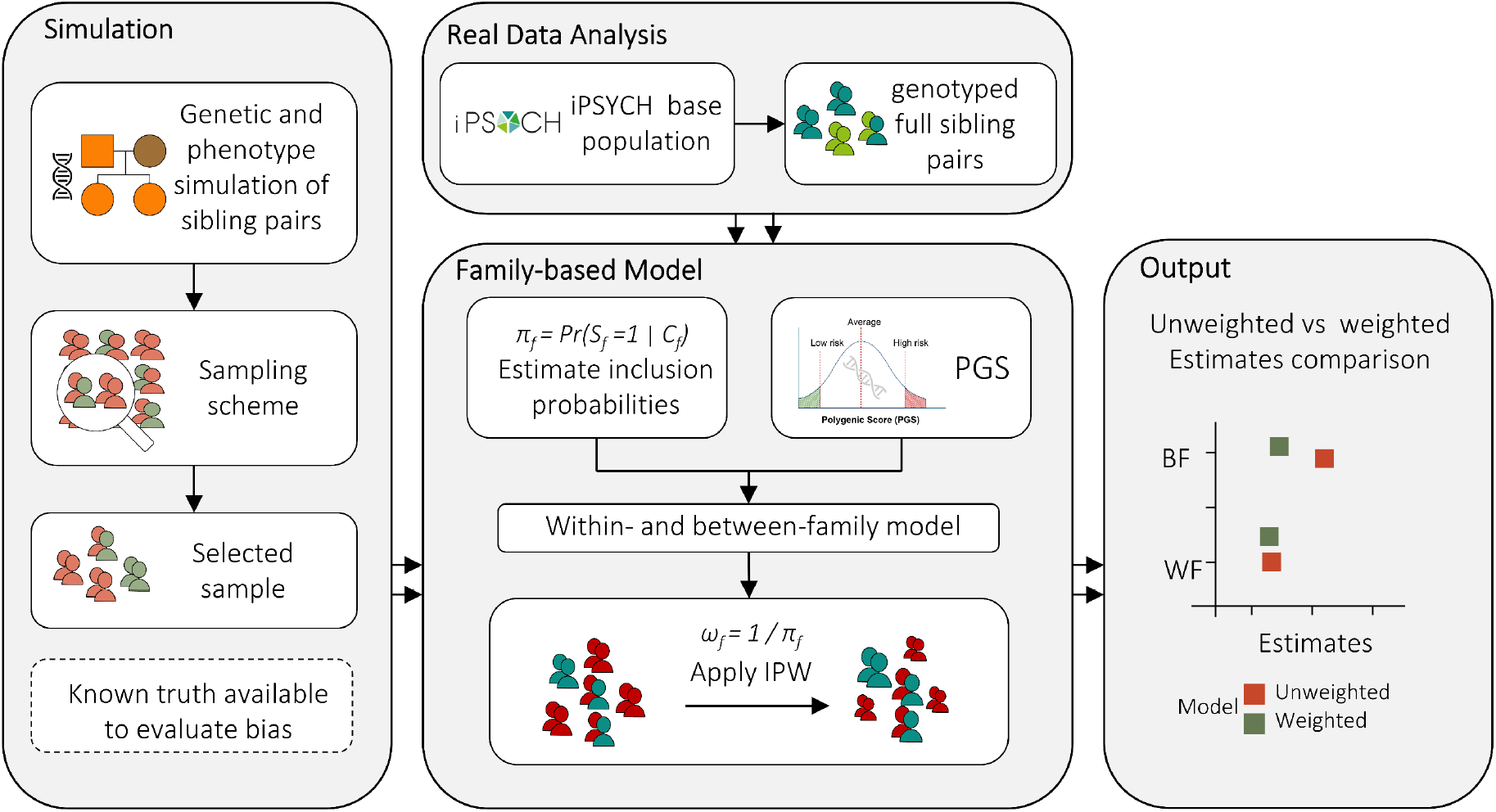
Overview of the study design and analytic workflow. π_f_ : Inclusion probability for family *f*: *S*_*f*_ : selection of family *f*; C_f_ : case configuration of family *f*; PGS: polygenic score; *ω*_*i*_ = sampling weight for pair *i*; IPW: inverse probability weighting; WF: within-family estimate and BF: between-family estimate.

### Simulation studies

To examine the effect of selection bias on within- and between-family polygenic score estimates, we simulated and analysed synthetic genotype and phenotype data. Genotypes of 10,000 independent SNPs were generated for 500,000 individuals (250,000 sibling pairs). A subset of 100 variants were assigned non-zero effects, and phenotypes were simulated under a liability threshold model assuming a total heritability of 0.6. A 5% prevalence threshold on the liability distribution was applied to define binary outcomes.

To evaluate how selection bias may distort PGS effect estimates, we introduced non-random sampling based on family case status. Families were stratified into three groups according to the number of affected siblings and were sampled at different rates to enrich the analytic sample for case-concordant sibling pairs. IPWs were then applied based on known sampling probabilities for each family stratum. We assessed the association between PGS and binary phenotypes using logistic regression models including within- and between-family PGS components as fixed effects. Models were fitted to the simulated population and biased sample, with and without IPW. Additional simulation scenarios with varying ratios of within- and between-family PGS effects were also considered. Details on simulations are described fully in the **Methods**.

Figure 2 displays the estimated within- and between-family effects of the PGS under different simulation scenarios. For each scenario, three models were fitted: using the full simulated dataset (unbiased reference), the biased unweighted subset, and the biased IPW weighted subset. Although the liability was modelled using linear combinations of within- and between-family PGS components with predefined coefficients (e.g., 0.5), all reported estimates correspond to log odds ratios from logistic regression on the resulting binary phenotype.

The positive control using full population data demonstrated the true parameter values were recovered across all conditions. In the biased subsets, unweighted estimates systematically deviated from the true effects, specifically demonstrating bias contamination in the between-family component. This distortion was more marked in scenarios under more case-enrichment extremes (top row). The application of IPWs substantially attenuated within-family as well as between-family bias, aligning the results more closely with those obtained from the full data. Across all settings, the weighted analyses consistently produced estimates with reduced bias compared to their unweighted fitted models.

### Application to real-world data

To evaluate the empirical relevance of selection bias in family-based genetic association studies, we applied the within- and between-family PGS model to genotyped full siblings from the iPSYCH cohort ^20^. The analytic sample included 7,404 full-sibling pairs with complete genotype and phenotype data. Analyses were conducted across six psychiatric phenotypes: attention-deficit/hyperactivity disorder (ADHD), autism spectrum disorder (ASD), major depressive disorder (MDD), bipolar disorder (BD), schizophrenia (SCZ) and anorexia nervosa (AN). Polygenic scores were constructed using GWAS summary statistics from independent external cohorts. Logistic regression models were fit for each trait, including both within- and between-family PGS components as predictors. To assess potential distortion due to sampling, all models were estimated with and without IPWs. Weights were derived from known inclusion probabilities of sibling pairs based on registry data. Further information on the sample demographics is presented in the **Methods** section.

As shown in Figure 3, we observed unweighted between-family estimates were consistently larger than corresponding within-family estimates across multiple phenotypes. This is consistent with findings from prior studies where between-family effects were interpreted as evidence of rGE. These effects, however, can result from many different types of confounding, e.g., population stratification, assortative mating ^10, 14^. Following the application of IPWs, several between-family estimates attenuated or shifted in direction. For example, in ASD, the between-family estimate decreased from 0.485 to 0.200, resulting in a smaller estimate than the corresponding within-family effect. A similar pattern was observed for MDD and AN.

**Figure 3.**
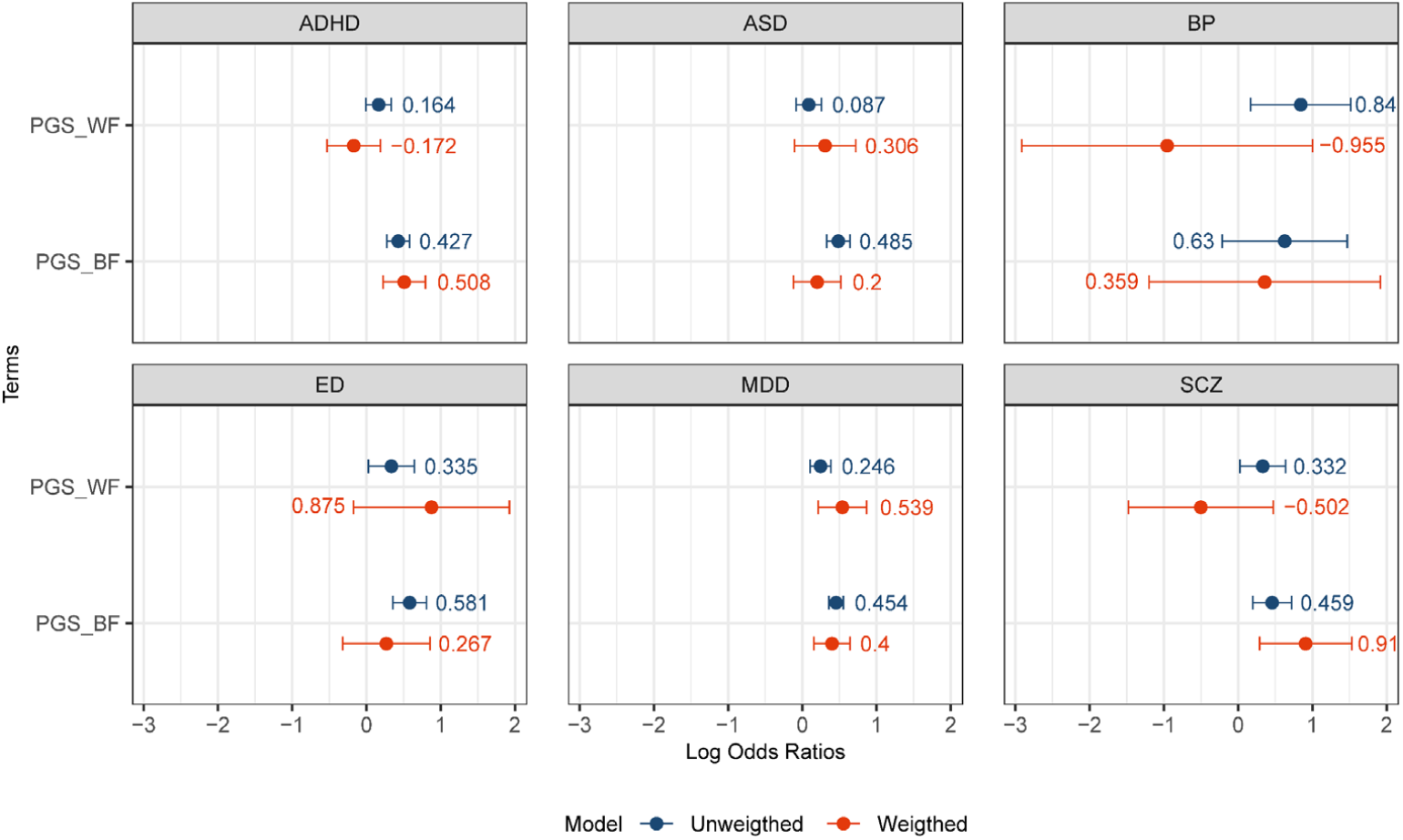
Estimated within-family and between-family effects of polygenic scores (PGSs) across six psychiatric phenotypes in the iPSYCH cohort. Each panel presents the log odds ratios from logistic regression models fitted with and without inverse probability weights (IPW). Blue points correspond to unweighted estimates, and red points to weighted estimates. Error bars indicate 95% confidence intervals.

### Prediction Accuracy

To assess whether decomposing the total PGS into within- and between-family components improves predictive performance, we conducted five-fold cross-validation using both simulated and real-world iPSYCH cohort data. Two models were compared: one including only the total PGS, and one including both within-family and between-family PGS components (See **Methods** for detailed information).

As shown in Figure 4, in the simulation analyses, predictive accuracy was evaluated using area under the curve (AUC) and observed scaled R^2^. Across all scenarios, the decomposed model (including within- and between-family components) and the total PGS model showed high discrimination, with AUC values in the range of 0.91–0.95. In scenarios with imbalanced effect sizes (e.g., WF=0.3, BF=0.7 or WF=0.7, BF=0.3), the decomposed model yielded slightly higher AUC and scaled R^2^ compared with the total PGS model, with average gains of ∼0.01–0.02 in AUC and modest increases in scaled R^2^. In more balanced scenarios (WF=0.5, BF=0.5), the two models performed similarly.

**Figure 4.**
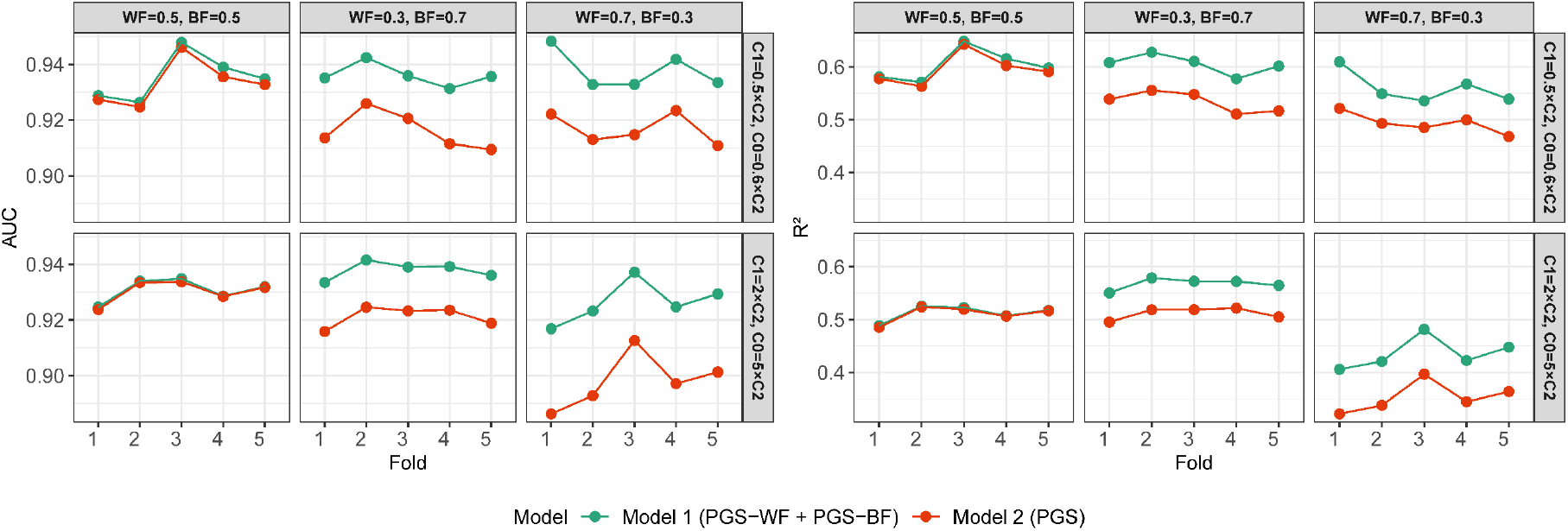
Prediction accuracy in simulated data. Comparison of predictive performance between the decomposed model (Model 1: within- and between-family PGS components) and the total PGS model (Model 2) across simulation scenarios. Panels show cross-validated area under the curve (AUC, right) and observed scaled R^2^ (left) for each scenario, defined by different ratios of within-family (WF) and between-family (BF) effects and by sampling probabilities. The top and bottom in each panel represent two biased sampling designs with varying inclusion probabilities for families with two cases (C2), one case (C1), and no cases (C0). Boxes represent the distribution of estimates across five folds.

In the real trait analysis (Supplementary Figure S1), conducted across six psychiatric phenotypes, the mean AUC difference between the two models ranged from –0.00093 to 0.007. Differences in observed scaled R^2^ showed a similar pattern, with only minor variation between specifications. No consistent improvement in prediction accuracy was observed.

## Discussion

This study examined the impact of selection bias on PGS estimates in family-based genetic association studies. We examined models that separate genetic effects into within-family and between-family components and evaluated the influence of biased sampling on these estimates. Using both simulated and real-world iPSYCH cohort data, we found non-random sampling can substantially distort between-family associations. Further, we demonstrated IPW, when based on known inclusion probabilities, can reduce this bias and yield more accurate estimates.

PGSs are increasingly being used for complex trait prediction. However, related mechanisms of the association of PGS and phenotypic variation remain inadequately explained. Family-based designs are often regarded as a robust method to addressing confounding resulting from shared environmental exposures or population stratification. Within-family models exploit random allele segregation conditional on shared parents, and shared sibling environment, thus constant within siblings, to estimate genetic effects while minimising confounding. Such comparisons enable the identification of potential mechanisms such as passive rGE,, where parental genetic liability is associated with the family environment provided to offspring, that may contribute to the predictive power of PGSs ^10, 14, 22^.

Although family-based designs are widely regarded as a robust method to partially mitigate rGE, they too are subject to methodological limitations. Research has highlighted within sibling analyses fixed-effects models can be biased in multiple ways ^23, 24^. Trejo et al., showed sibling analysis underestimates direct genetic effects within families, while potentially inflating between-family estimates ^24^. Furthermore, PGS studies remain vulnerable to selection bias. This is particularly relevant in large biobank studies, where participants and underlying population often differ systematically. For example, a recent investigation demonstrated individuals who enroll in volunteer-based cohorts tend to differ from patients in the underlying healthcare population with respect to demographic characteristics, clinical history, and healthcare utilisation. The authors demonstrated such differences can bias PGS estimates for psychiatric disorders ^17^. Furthermore, within-family models inherently have a reduced effective sample size compared with between-family analyses. Although multiple sibling pairs can be formed within a larger sibship, these pairings do not contribute independent information because siblings share a substantial proportion of their genotype and environment. As a result, estimates derived from within-family models may be less precise, and the ability to detect modest genetic effects is correspondingly reduced. An additional consideration in family-based analyses is that sibling participation itself may be non-random. In cohorts where families have more than two children, it is common for only a subset of siblings to be genotyped. If participation is related to traits that correlate with the PGS, for example, behavioural, clinical, or sociodemographic characteristics shared within families, this may introduce bias in within-family estimates, as the observed sibling pairs are not representative of all possible sibships.

IPW weights individuals by the inverse of their estimated probability of inclusion, aiming to restore population representativeness in the study sample ^25^. Lee et al., in a psychiatric genetics study using hospital-based biobank data, found unweighted models tend to yield larger genetic association estimates compared to IPW-adjusted models ^17^. This highlights the importance of accounting for sampling structure when estimating PGS predictive performance and penetrance in clinical and epidemiological studies. However, reliable sampling weights are not always available, particularly in cohorts lacking linkage to population registers or detailed information on non-participants, which limits the broader applicability of this correction.

In our simulation studies we demonstrated selecting on case status alters the distribution of the outcome in the sample and introduces systematic bias in PGS effect estimates. Between-family estimates were particularly sensitive to this form of selection, with increasing deviation from the true effects found under more extreme sampling schemes. In contrast, within-family estimates remained relatively stable across all scenarios. When IPWs were applied to correct for known sampling probabilities, both within- and between-family estimates converged towards their true values. This demonstrates IPW can mitigate bias within family-based models, particularly with complex data or non-representative samples.

In the application to the iPSYCH cohort, we observed a consistent pattern in which unweighted between-family estimates exceeded within-family estimates for several psychiatric phenotypes, such as ASD and MDD. Previously this was assumed to be indicative of rGE, whereby genetic predisposition is correlated with environmental exposures that also influence the phenotype. Several studies, including those for educational attainment, intelligence, and psychiatric traits reported similar findings ^10, 14^. However, these differences may also result from selection processes that enrich the sample for specific combinations of genotypes and outcomes. Our analysis demonstrated IPWs, derived from empirically estimated inclusion probabilities, attenuated several between-family effect estimates. In such conditions, for example, ASD and MDD, between-family estimates fell below their corresponding within-family estimates in magnitude. These findings may reflect if family inclusion probability depends on genotype or phenotype-related characteristics, residual bias may remain in the resulting population estimate even after controlling shared environmental influences.

We also compared the predictive performance of models using either total PGS or separate within- and between-family components. In simulations where within- and between-family effects were equal, predictive accuracy was comparable. However, when these components contributed unequally to the phenotype, models including both effects had superior AUCs. In contrast, analyses of real-world data showed minimal differences in AUC between models. While model decomposition may enhance prediction in some simulated settings, its added value in real-world data may be limited, likely due to modest effect sizes, shared confounding, and low signal-to-noise ratios. Nonetheless, separating PGS components remains useful to clarifying sources of association and assess robustness to bias.

Our study provides empirical support for the use of IPW as a selection bias correction method, particularly in the absence of reliable sampling probability. However, this approach is not without limitations. A key challenge remains in applying this framework to large population-based biobanks lacking administrative register links. Cohorts such as the UK Biobank are affected by well-documented selection biases, such as healthy volunteer bias and differential participation based on socioeconomic or health-related factors ^26-29^. It may be possible to approximate inclusion probabilities through a combination of expert knowledge and data-driven methods. Previous studies have shown the importance of integrating clinical data with variable selection techniques informed by high-dimensional data, notably when working with electronic health records ^30-33^. While such strategies hold promise, their effectiveness depends heavily on the availability of representative data, and the applicability of the model to the target cohort. As healthcare systems differ in structure and population coverage, selection models trained in one context may not generalise readily to others. Therefore, efforts to adjust for selection bias in large-scale genomic studies will likely require context-specific modelling approaches to reflect the unique composition and dataset specific selection patterns. Furthermore, if significant predictors of inclusion are unmeasured or poorly captured, residual selection bias may persist. Within the present study simulations were necessarily simplified, thus may not fully capture the complexity of real-world genetic and environmental structures. Finally, while the inclusion probabilities for the iPSYCH cohort were derived from well-documented sampling schemes, small inaccuracies in their estimation may still influence IPW performance. The generalisability of these findings to other populations, traits, and study designs should be further explored in future work.

## Conclusion

This study demonstrates that selection bias is a quantifiable source of distortion in family-based PGS analyses. While family-based study designs address many forms of confounding, they do not eliminate the impact of non-random sampling. Effect sizes from studies that do not adjust for selection should be interpreted cautiously. Applying IPWs based on empirically estimated inclusion probabilities demonstrates the potential to improve estimate accuracy. Our findings highlight the need for careful consideration of sampling processes in genetic studies and demonstrate the value of combining family-based designs and methods to account for selection. Ultimately, observational cohorts are essential resources to study the causes and consequences of health and disease. Ensuring accurate sample representation is critical for the translation of research findings into healthcare advances to maximise population benefit. Thus, efforts to improve representativeness and methods to assessing the impact of selection are essential.

## Method

### Statistical model

To estimate the association between PGS and the binary phenotype while accounting for familial structure, we applied a generalised linear mixed-effects model to partition the total genetic association into within- and between-family components. This allows for the separation of direct genetic effects and potential confounding from shared family-level influences. The model includes both individual deviation from the family mean PGS (within-family component) and family average PGS (between-family component) as fixed effects. Let *Y*_*ij*_ denote the binary outcome for individual *i* in family *j*, and *PGS*_*ij*_ represent the individual’s PGS. The model is specified as:

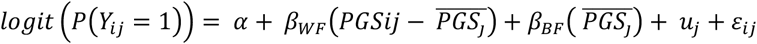

where 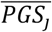 is the mean PGS for family *j, β*_*WF*_ the within-family effect of PGS, and *β*_*BF*_ the between-family effect. A family random intercept 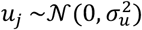 accounts for unmeasured familial influence shared by siblings. *α* denotes the intercept, and the residuals are represented by *ε*_*ij*_, with 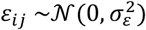. We use and expand this model with various fixed effects to separate the association between PGS and phenotype into within family (individual level) and between families effects. To assess the influence of selection bias on effect estimates, we applied IPW with weights derived from estimated inclusion probabilities. Separate models were fit with and without weighting, and comparisons were used to evaluate the impact of biased sampling on the estimates. Covariates for sex and age were included in all models but are omitted from equations for brevity.

### Simulation

We simulated a dataset of 500,000 individuals, grouped into 250,000 full-sibling pairs, with 10,000 bi-allelic single nucleotide polymorphisms (SNPs). Genotype data were generated using LDAK ^34^. Simulated SNPs followed Hardy–Weinberg equilibrium with minor allele frequencies (MAF) drawn uniformly from 0.05 to 0.5.

Phenotypes were simulated under a liability threshold model, with assumed heritability of 0.6. A total of 100 SNPs were designated as causal variants, with corresponding effect sizes sampled to reflect additive genetic contributions. A continuous liability trait was generated for each individual, summing the weighted genotype dosages across the causal variants and adding normally distributed environmental noise. The variance of the environmental component was scaled to achieve the target heritability on the liability scale. A binary phenotype was then defined by applying a prevalence threshold of 5%, with cases identified as individuals in the upper tail of the liability distribution.

PGSs were computed for all individuals using simulated effect sizes. Each score was calculated as the sum of genotype dosages weighted by their corresponding SNP effects. For each sibling pair, the PGS was decomposed into a between-family component, defined as the family mean, and a within-family component, defined as the deviation from the family mean. The liability was modelled as a linear function of both within- and between-family PGS components. To evaluate their relative contributions, we simulated scenarios with varying ratios of the two components. In the first scenario, both components contributed equally to the liability, such that the standardised effects of the within-family and between-family components were set to 0.5. In the second scenario, the within-family component had a larger contribution and was set to 0.7 for the within-family and 0.3 for the between-family component. In the third scenario, the relative contributions were inverted, assigning a larger standardised effect to the between-family component (0.7) and a smaller effect to the within-family component (0.3).

To introduce selection bias, families were classified into three strata based on the number of affected siblings: those in which both siblings were cases (C2), one sibling was a case (C1), or neither sibling was affected (C0). All C2 families were retained. From the remaining families, two sampling schemes were applied to reflect varying degrees of case enrichment and replicate real-world ascertainment patterns. In one scenario, 50 % of C1 families and 60% of C0 families were sampled relative to the number of C2 families. In a second scenario, C1 and C0 families were sampled at rates equivalent to twice and five times the number of C2 families, respectively. This sampling scheme was designed to approximate enrichment of case-control sibling pairs as observed in real-world data and described below. IPWs were computed from sampling probabilities assigned to each family stratum.

Analyses were conducted both with and without the application of IPWs. Although the data structure consisted of clustered sibling pairs, random intercepts were not included in the simulation models to maintain analytical simplicity and isolate the fixed effects of interest.

### Prediction accuracy

To evaluate and compare the predictive performance of models using total PGS and decomposed within- and between-family PGS components, we conducted five-fold cross-validation. To ensure independence of observations across folds, sibling pairs were kept together by assigning the same fold identifier to each family. Unique family identifiers were randomly allocated to one of five folds. For each fold, a logistic regression model was trained on the remaining four folds and tested on the held-out fold. Two models were fitted: one including the total PGS as a predictor, and one including both the within-family and between-family components. For each model and fold, predictive performance was assessed using the AUC and observed scaled R^2^.

### Study population

This study utilised data from the population based iPSYCH2015 case-cohort sample. This cohort has been described in detail elsewhere ^20^. In brief, iPSYCH2015 is an extension of the original iPSYCH2012 sample ^35^, drawn from a source population of singleton births in Denmark between 1 May 1981 and 31 December 2005. Inclusion criteria required individuals to have known mother and to be alive and residing in Denmark at age one. This cohort includes the individuals diagnosed with major psychiatric disorders (including ADHD, ASD, BP, MDD or SCZ), identified via ICD-10 codes ^36^ from the Danish Psychiatric Central Research Register ^37^. It also included the AN samples from the Anorexia Nervosa Genetics Initiative (ANGI) ^38^ and Eating Disorder Genetic Initiative (EDGI) ^39^, as they were samples within the same framework as iPSYCH2015. It further includes a randomly selected population based subcohort of approximately 51,000 individuals, that are broadly representative of the Danish population. Full sibling pairs were identified using the register data. Pairs were defined as full siblings if they shared both registered parents and met iPSYCH inclusion criteria. The final analytic dataset included 7,404 unique sibling pairs with complete genotype and covariate data. For each trait, the number of sibling pairs with non-missing phenotype data was as follows: 4,710 for ADHD, 4,099 for ASD, 1,910 for AN, 2,028 for SCZ, 1,608 for BP, and 5,207 for MDD.

### Polygenic score (PGS)

PGSs were constructed as the weighted sum of trait-associated alleles, with weights derived from SNP effect sizes reported in GWASs. Summary statistics were obtained from the largest available GWAS for key psychiatric and developmental phenotypes, including ADHD ^40^, AN ^41^, ASD ^42^, SCZ, BP and MDD ^43^. We generated the PGS using the LDpred2 software ^44^ and excluding iPSYCH samples.

### Inclusion probabilities and sampling weights

The iPSYCH cohort consists of two population-based case-cohort samples drawn from Danish national registers. The first sample was established in 2012. From this source population, all individuals diagnosed with at least one listed major psychiatric disorder by the end of 2012 were included and a random sample comprising approximately 2.037% of individuals was selected as a subcohort. The second sample was constructed in 2015 from an extended source population comprising all individuals born up to 31 December 2008. This sample included all incident cases diagnosed by the end of 2015 and a 1.267% random sample from the population.

Consequently, the probability of inclusion for subcohort members varies by birth cohort and sampling phase. For individuals born between 1981 and 2005, the inclusion probability was estimated at 3.278%. This value was derived under the assumption individuals could have been selected in either of the two iPSYCH sampling phases. For individuals born between 2006 and 2008, the inclusion probability was 1.267%, corresponding to the second sampling phase ^21^.

Let *π*_*i*_ denote the probability that individual *i* was included in the genotyped cohort. Based on the design above,

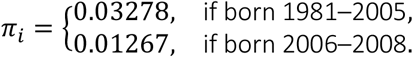

As the sibling pairs used in the real-world analyses were drawn from the sampled sibling pairs in the iPSYCH cohort, it was necessary to determine the sampling probabilities for sibling pairs rather than individuals. Under the assumption of independent selection, the inclusion probability for a given pair can be calculated as the product of the individual inclusion probabilities of the two siblings. For siblings *i* and *j*, the pair-level probability was therefore defined as

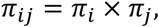

and the inverse probability weight assigned to the pair was

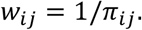

However, we considered the possibility that selection might not have been completely independent. For instance, if clustering mechanisms or dependencies existed such that the selection of one sibling influenced the inclusion probability of the other, the assumption of independence is violated. To assess this, we examined the structure of the original cohort using nationwide register data. We identified all possible full sibling pairs and stratified them by case status. For each stratum, we computed the expected number of sibling pairs included under the assumption of independent sampling. Formally, for each stratum *s* ∈ { *C*2, *C*1, *C*0}, the expected count was

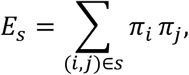

and this was compared with the observed number of genotyped pairs *O*_*s*_. These expected counts were compared with the observed number of genotyped sibling pairs within iPSYCH. The observed and expected frequencies of sibling pairs across all case-status strata were closely aligned, with minimal discrepancies. This indicates the independence assumption holds to a reasonable degree. Therefore, we proceeded by estimating sibling pair inclusion probabilities as the product of the individual-level inclusion probabilities of the two siblings.

The resulting weights *w*_*ij*_ were incorporated into the within-family logistic regression models so that each sibling pair contributed to the likelihood in proportion to the inverse of its probability of selection, aiming to recover estimates representative of the underlying population.

## Data Availability

Data are not publicly available due to Danish data protection legislation and restrictions governing access to individual-level genetic and health register data. Access requires approval from the relevant Danish authorities and can be obtained only through an approved research collaboration

